# Optimal annual COVID-19 vaccine boosting dates following previous booster vaccination or breakthrough infection

**DOI:** 10.1101/2024.07.25.24311016

**Authors:** Jeffrey P. Townsend, Hayley B. Hassler, Alex Dornburg

## Abstract

COVID-19 booster vaccinations mitigate transmission and reduce the morbidity and mortality associated with infection. However, the optimal date for booster administration remains uncertain. Geographic variation in infection rates throughout the year make it challenging to intuit the best yearly booster administration date to effectively prevent infection, and also challenging to provide best guidance in how to alter booster administration in response to a breakthrough infection. Therefore, we leverage longitudinal antibody and reinfection probabilities with spatiotemporal projections of COVID-19 incidence to develop a geographically-informed approach to optimizing the timing of booster vaccination. Additionally, we assess the delay in booster vaccination that is warranted following breakthrough infections whenever they occur during the year, enabling an approach that acknowledges and respects diverse immune statuses, thereby addressing a substantial barrier to uptake. Our results provide guidance for individual decision-making and healthcare provider recommendations, as well as optimal public health policies.

**Significance Statement:** COVID-19 booster vaccinations are pivotal in reducing disease transmission. However, optimal schedules that would most successfully mitigate adverse health outcomes have not been rigorously determined. Spatial and temporal surges of infections, including breakthrough infections, challenge the implementation of effective boosting strategies. We leverage antibody data and incidence projections to develop a geographically-informed schedule for yearly booster administration and quantify appropriate delays in booster vaccination post-breakthrough infection, thereby accounting for immune status and enhancing vaccination inclusivity. Our findings offer crucial information for individual decision making, healthcare provider guidance, and policy aimed at optimizing the impact of booster vaccination on public health.

## Introduction

COVID-19 vaccines confer protection against infection (1) and severe disease (2). For example, in New York City alone, vaccination prevented nearly 300,000 cases, 50,000 hospitalizations, and 8,500 deaths between December 14, 2020 and July 15, 2021 (3). To provide long-term protection, booster vaccination is necessary, which addresses the rapid waning of SARS-CoV-2 antibody levels (4, 5). However, the date when it is optimal to administer yearly booster vaccinations is not clear. This decision is complicated by the fact that SARS-CoV-2, like other coronaviruses, exhibits substantial spatiotemporal variation in incidence (6, 7). Aligning booster vaccination timing so that it provides peak protection during peak incidence may be crucial to maximizing their benefit.

In addition to considering the optimal dates for yearly booster vaccination, it behooves us to evaluate whether and for what duration it would be appropriate to delay that date in the case of breakthrough infection. Following infection and an immune response against SARS-CoV-2 in a healthy individual, the resulting production of antibodies would offset benefits of an immediate booster vaccination (8). Consequently current guidance suggests an available updated booster might beneficially be delayed to reinforce immunity later in the year, with the U.S. Centers for Disease Control currently suggesting a delay of up to three months (9). However, this guidance has not been rigorously evaluated. The lack of rigorous guidance has created a significant barrier to vaccine uptake. Therefore, it is vital to establish policies that integrate the timing of booster vaccinations with an awareness of breakthrough infections to minimize risk.

The question of when to boost has been extensively researched for influenza, where many years of endemic seasonal incidence data have been collected. Influenza exhibits seasonal infection surges associated with climate (10) and analyses of these data have provided expert guidance regarding optimal timing of influenza vaccination (11–13). Consequently, in the United States, the Center for Disease Control has recommended influenza booster vaccination between September and October (14). Indeed, in countries that span temperate and tropical latitudes such as China, the optimal timing of yearly influenza vaccination can vary within the nation by as much as several months (15). These findings regarding influenza—a common, seasonal respiratory virus—support performing similar analyses regarding the spatiotemporal optimality of COVID-19 booster vaccination. However, there are far fewer years of seasonal infection prevalence data on which to base predictions. Moreover, COVID-19 infections during these few years do not reflect endemic seasonality, and instead have been characterized by pandemic surges driven by historical contingencies, early saltations of viral evolution, and wholly naive versus differentially exposed immunological states of populations. Therefore, an alternate approach to evaluating optimal timing is necessary until there is sufficient accumulation of long-term empirical data on seasonality.

Such an alternate approach has been enabled by recent research that projected the seasonality of endemic COVID-19 based on ample coronavirus incidence data, establishing clear differences in seasonal circulation with substantial spatiotemporal heterogeneity in peak incidence across the northern hemisphere (16). These results, generally parallel typical respiratory virus incidence patterns, featuring peaks of “flu and cold season” with some specificity in each locality. However, in isolation these results give no direct guidance regarding when to administer booster vaccinations. The optimal timing of booster vaccinations depends jointly on the risk of infection due to seasonality and on the long-term waning of protection subsequent to booster vaccination (17). Rigorous estimates of long-term protection subsequent to booster vaccination have been obtained by leveraging longitudinal antibody and reinfection data from the close evolutionary relatives of SARS-CoV-2 as well as infection and antibody data on SARS-CoV-2 to estimate the durability of immunity following natural infection (18), primary vaccination (19), and boosting (20, 21). These analyses—subsequently validated by comparison to empirical data (22–24)—indicate that statistical approaches derived from evolutionary medicine can illuminate reinfection risks with high accuracy and precision. Yet the impact on yearly infection of administering yearly booster vaccinations at specific times in specific locations remains unknown—as does the impact of breakthrough infections at specific times of the year on the optimal timing of booster vaccination.

Here we integrate the waxing probabilities of infection subsequent to antigen exposure with projections of the expected seasonal variation in frequency of infection for endemic SARS-CoV-2 (16), to perform a high-resolution investigation of prospective timings of booster vaccinations that maximally curtail infection. We evaluate the optimal boosting date over the year for individuals who have not been infected over the previous year. Then we analyze similar optimal timings for individuals who have been infected during that year to determine any advantage to delaying the booster, depending on the date of infection. Such knowledge of optimal booster vaccination timing will be helpful for physician and individual decision-making in the context of ongoing endemic disease, and is crucial for effective vaccination policy that suppresses morbidity and mortality as a consequence of COVID-19.

## Materials and Methods

We quantified relative monthly probabilities of infection based on seasonal incidence predictions for endemic COVID-19 (16) for 12 locations within the Northern Hemisphere: (1) Rochester, MN USA; (2) New York City, NY USA; (3) Edinburgh, UK; (4) Stockholm, Sweden; (5) Trøndelag, Norway; (6) Gothenburg, Sweden; (7) Amsterdam, Netherlands; (8) South Korea (nationwide); (9) Yamagata, Japan; (10) Guangzhou, China; (11) Sarlahi, Nepal; and (12) Beersheba, Israel. These predicted incidences were derived by employing a phylogenetic ancestral and descendant state approach that leveraged long-term data on the incidence of circulating HCoV coronaviruses in diverse geographic areas (16). These projections were robust to the choice of the model of trait evolution as well as the choice of molecular trees, relative phylogenetic chronograms, and non-recombinant alignments.

These seasonal incidence predictions are derived from long-term endemic coronavirus prevalences with no vaccinations or other interventions. However, an individual recently vaccinated against SARS-CoV-2 will experience lower risk of infection than an unvaccinated individual in an endemic scenario with no interventions. Therefore, for each date of booster vaccination, over the first month following the date of booster vaccination, we reduced the seasonality-based probability of infection by the daily proportional protection demonstrated in Pfizer-BioNtech booster (BNT162b2) vaccination clinical trials against the delta variant (B.1.617.2) of SARS-CoV-2 following two primary vaccinations (25). This vaccine-based daily probability of infection was calculated via linear interpolation between the probability of no breakthrough infection at clinical trial sampling times post booster vaccination (one week and two weeks), at peak vaccination at one month, and at one year post-vaccination. For the second through 12^th^ months following the booster vaccination date, reductions of the seasonal probability of infection across the rest of the year due to booster vaccination were quantified by analysis of antibody waning and corresponding infection probability (21). Antibody waning data was associated with Pfizer-Biontech COVID-19 vaccination and SARS-CoV-2 infection, and infection probabilities were associated with antibody levels based on an ancestral and descendent states analysis of long-term antibody data on healthy individuals who experienced other human-infecting coronaviruses (HCoVs, 18). This analysis incorporated construction of a maximum-likelihood molecular phylogeny of human-infecting coronaviruses. This phylogeny enabled comparative analyses of peak-normalized optical density levels of blood-based IgG antibodies to nucleocapsid protein, spike protein, and whole-virus lysate, that were coupled with reinfection data on endemic human-infecting coronaviruses. Expected declines in antibody levels over time as well as the probabilities of reinfection based on antibody level were estimated from empirical data supplemented by ancestral and descendent states analyses. The resulting probabilities of reinfection provided times to reinfection after recovery under conditions of endemic transmission for SARS-CoV-2. The antibody waning profiles and infection probabilities from this approach have proven consistent with multiple time points reported in subsequent but shorter-term wholly empirical studies (22–24).

For each day of the year, we calculated the cumulative yearly probability of infection, wherein the probability of infection each day was equal to the probability of not being infected each previous day times the probability of being infected on the day of interest. For each day of the year, we then plotted these cumulative yearly probabilities of infection. The booster vaccination date on which the cumulative yearly probability of infection is at its lowest represents the optimal yearly booster vaccination date.

To visualize how shifts in seasonal incidence influence the optimal timing of booster vaccination, we plotted probability mass functions of infection by month in the 12 locations examined. Incidence bars were colored to indicate the monthly probabilities of infection for an individual who (1) is booster vaccinated over a period of three years on the optimal yearly booster vaccination days, and (2) is booster vaccinated once, then foregoes booster vaccination on the subsequent two optimal yearly booster vaccination days.

We also examined the delay of booster vaccination that should be recommended in the case of a breakthrough SARS-CoV-2 infection. We modified our approach above to address breakthrough infections occurring on each date between optimal yearly booster vaccinations by renewing protection at the point of breakthrough infection. From peak antibody response to breakthrough infection at one month following the onset of infection, antibody level followed the waning function of Townsend et al. (21). During the first month after breakthrough infection, reductions of the probabilities of breakthrough infection were set to be consistent with observations for the BNT162b2 booster clinical trial versus the B.1.617.2 variant. Optimality of the delayed date of booster administration was then evaluated as the cumulative probability of infection spanning from the yearly optimal booster vaccination date following the infection through to the following yearly optimal booster vaccination date.

## Results

Studies that have not incorporated the date of boosting into their analyses have found that annual boosters reduce the probability of infection by about 67% (19), suggesting yearly booster vaccination on any date is beneficial to prevention of infection. Analysis incorporating seasonality as well as the date of booster vaccination reveals substantial additional variance in efficacy by date (**Fig. 1**). The benefit of optimally timing booster vaccination is modulated by the seasonality of disease at each geographic location. In general, each location exhibits an optimal date on which boosting offers a 3–4-fold increase in protection from infection over other times of the year, and a range of nearby dates that feature similar benefits. The timing of these periods varies substantially between locations. In New York, USA (**Fig. 1A**), yearly booster vaccination on September 15^th^ provides the lowest yearly probability of infection. This benefit diminishes slightly with delay as the yearly probability of infection increases to a 3.6-fold higher booster vaccination administration on the least effective date, January 24^th^. Yearly booster vaccination dates later in the year than this least effective date yield increasing benefits up to the optimal date.

**Figure 1.**
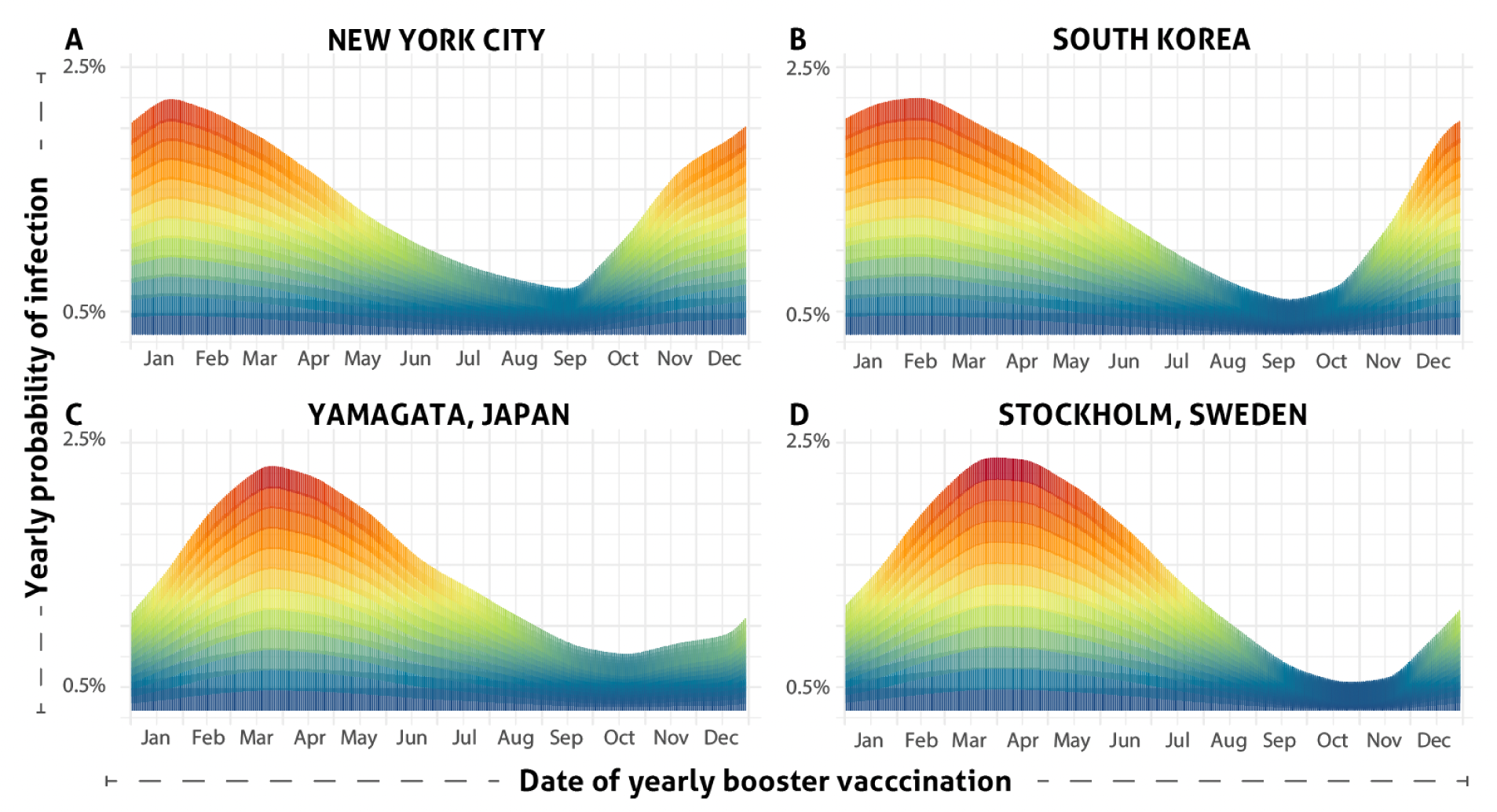
Yearly probability of infection (red: high to blue: low) based on yearly date of booster vaccination in (**A**) New York, (**B**) Stockholm (**C**) Yamagata, and (**D**) South Korea.

A similar pattern to that of New York is predicted for South Korea (**Fig. 1B**), where the date with the lowest yearly probability of infection is on September 19^th^. Boosting in September in South Korea is predicted to confer a nearly 5-fold increase in protection relative to delaying boosting to January or February. For Yamagata Japan (**Fig. 1C**), the yearly booster vaccination date with the lowest probability of infection was slightly later, on October 18^th^. booster vaccination on dates later than optimal result in a less steep rise in probability of infection compared to South Korea, but do obtain a nearly 5-fold differential in yearly infection probability. Yearly probabilities of infection in Stockholm, Sweden (**Fig. 1D**) were similar across the year to those for Yamagata, supporting booster vaccination in October. In Stockholm, infection risks remain similar for yearly booster vaccination in November, then rapidly rise to a nearly 5-fold differential compared to a much less effective yearly booster vaccination administered in the spring.

For other locations in the Northern Hemisphere, probabilities of infection throughout the year were often similar, reflecting a strong relationship that supports booster vaccine administration on a date that minimizes infection risk by preceding anticipated surges in SARS-CoV-2 incidence (**Fig. 2**). On average across locales, the optimal yearly date of booster vaccine administration preceded the highest anticipated prevalence by 2.7 months (95% CI: 1.9–3.4; **Supplementary Fig. 1**).

**Figure 2.**
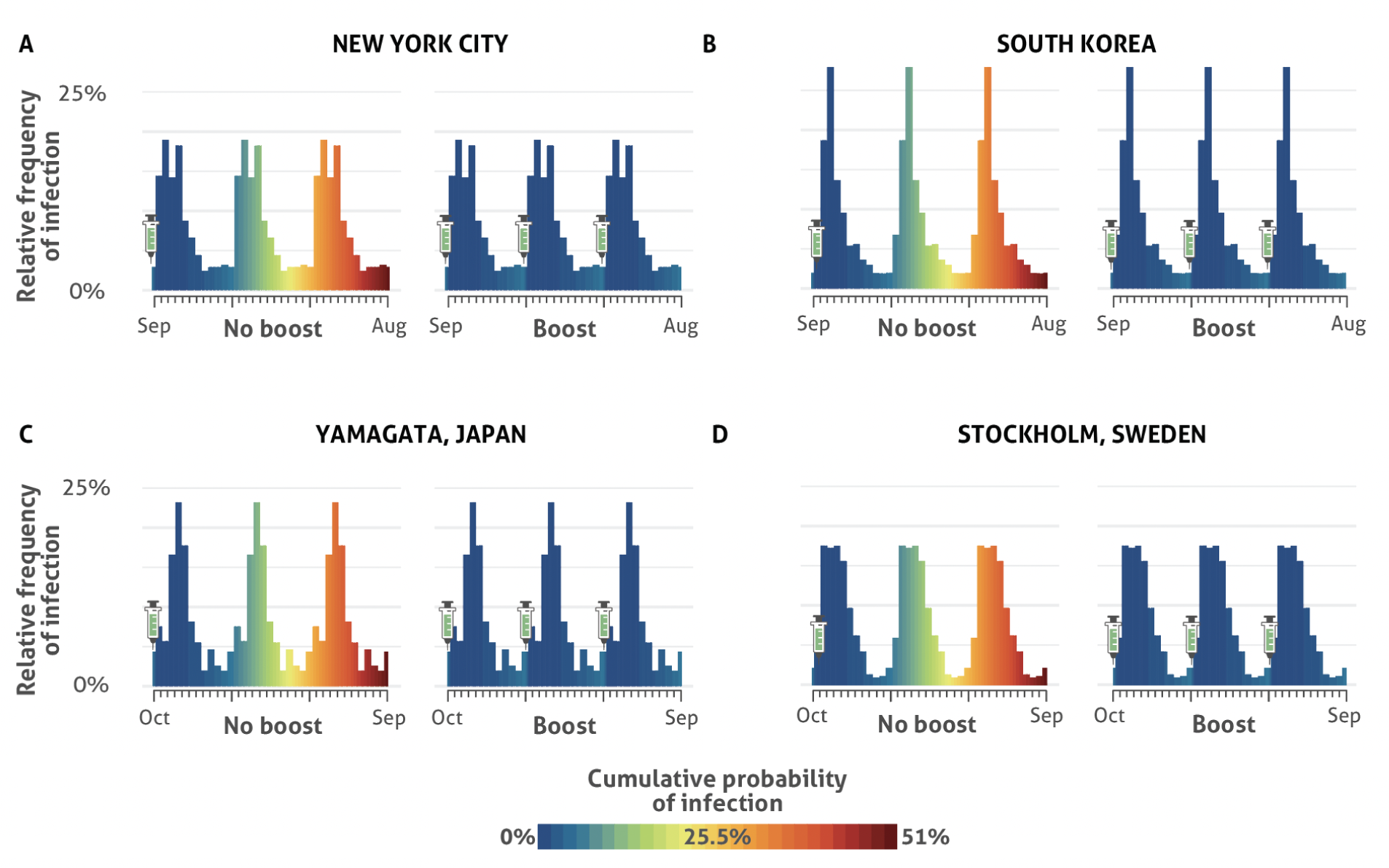
Cumulative probabilities of infection—indicated by the blue / green / yellow / orange / red color gradient—in the year after booster vaccination on the optimal date and in the two following years without the yearly booster vaccination, in (**A**) New York, (**B**) Stockholm, (**C**) Yamagata, and (**D**) South Korea. The heights of the bars reflect projections of seasonal population-level incidence patterns under endemic conditions without incorporating a population-wide effect of booster vaccination, i.e. assuming that low yearly global uptake (26) remains unchanged.

The optimal timing of booster vaccination can be substantially altered by a breakthrough infection that occurs during the interval between yearly optimal booster vaccination dates (**Fig. 3**). As might be expected, an infection occurring just before the yearly optimal booster vaccination date leads to an alternate date that would be optimal for that individual for that year. For instance, in New York, a breakthrough infection occurring on any date between September through the beginning of February has practical equivalence of benefit to booster vaccination on the optimal yearly date of September 15^th^ (**Fig. 3A**). This equivalence includes rare individuals who are exposed to a sufficient viral dose shortly after boosting: these individuals are not expected to be in a substantially different situation than someone who was only vaccinated with a booster, and likely should receive booster vaccination on or near its next availability, i.e. on the optimal yearly date of September 15^th^. Breakthrough infection occurring in the middle of March provides immunity such that the benefit booster vaccination is roughly equivalent anywhere along the span through to the following January (**Fig. 3A**). For an individual infected in August—just before what would typically be the yearly optimal booster vaccination on September 15^th^—the next booster vaccination should be delayed and occur some time between December and the following June.

**Figure 3.**
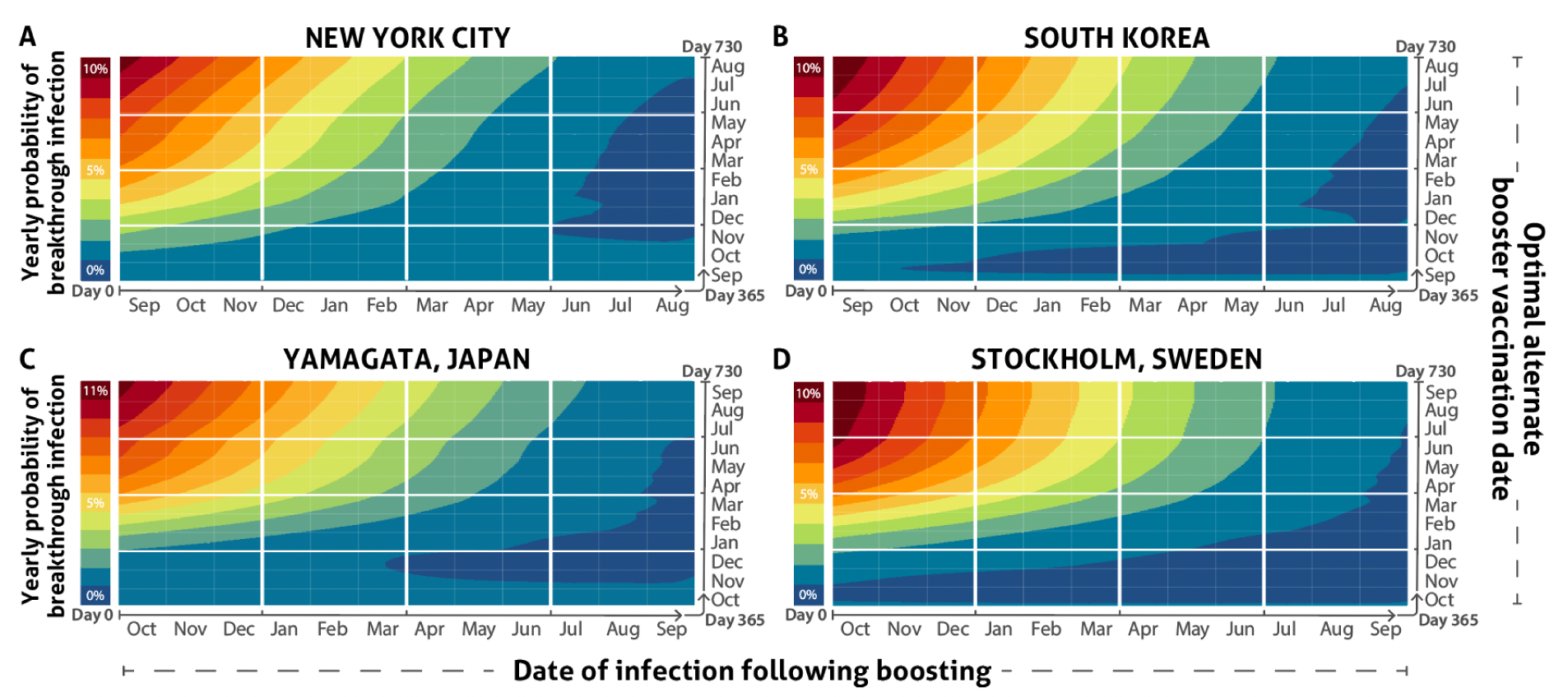
Probability of infection upon delaying booster vaccination to each date of the year, in response to a breakthrough infection occurring in the interval between optimal booster vaccination dates in (**A**) New York, (**B**) Stockholm (**C**) Yamagata, and (**D**) South Korea. Day 365 is the estimated optimal yearly booster vaccination date. Infection will delay the optimal booster vaccination date beyond day 365 to a greater and greater degree as the interval between the yearly optimal booster date and the date of infection increases.

Optimal booster timing was similarly delayed by breakthrough infection in other geographic locations (**Fig. 3B–D**), and similarly varied depending on infection date. Generally, individuals experiencing infection just subsequent to the optimal yearly booster vaccination date would benefit most by continuing to vaccinate on the yearly optimal date for each location. In contrast, those infected six months after the optimal yearly booster vaccination date will benefit most in terms of probability of infection over a yearlong span by delaying receipt of the next annual booster by several months. For individuals who were infected just prior to the next optimal yearly booster vaccination date, the optimal delays of booster vaccination exceeded nine months for nearly all locations. Analyses of additional geographic areas revealed that in some geographies in which anticipated seasonal infection trends are more muted, there was less pronounced benefit to optimizing booster vaccination timing (**Supplementary Fig. 2**).

## Discussion

Here we have shown that the optimal timing of yearly booster vaccination in the temperate Northern Hemisphere is typically in the autumn to early winter, with moderate variance in timing across locations. Accounting for annual syncopations of incidence represents an opportunity to increase the utility of booster vaccinations. Our analyses suggest that individuals who experience breakthrough infections with access to only one booster vaccination per year can beneficially delay their upcoming autumn booster later than the yearly optimal booster vaccination date. The amount of delay that provides the greatest benefit of booster vaccination is contingent on when the breakthrough infection occurs. Minimal delay in the next booster vaccination is warranted if an infection occurs shortly after a booster vaccination on the yearly optimal date. A breakthrough infection that occurs six to twelve months after booster vaccination warrants a delay of the booster vaccination by a comparable number of months. Identification and communication of appropriate booster timing to public health authorities, vaccine providers, and recipients along with guidance as to how to adapt that timing in response to infection enables more effective booster vaccinations that are cognizant of yearly incidence and infection history and therefore more universally inclusive in nature.

Our yearly optimal booster vaccination dates are based on endemic seasonality of COVID-19 infection. However, infection trends for the near-term may not adhere to endemic seasonality and may be subject to greater variability of surge timing and intensity. Moreover, our analysis assumes that the typical endemic seasonality is applicable each year. Seasons of endemic respiratory viruses are known to vary in severity as well as to surge early or late in some years (27–29). Indeed, in years where the seasonal surge starts early, booster vaccination will likely serve to prevent infection best if performed earlier than the optimal date indicated in this study.

Across locations, the intervals between the optimal yearly booster vaccination dates and the peaks of monthly incidence were fairly consistent from location to location. This geographical consistency implies that a surge occurring earlier than usual at a location should be addressed by advancing the dates of booster administration in that location proportionately. Similarly, in years where the seasonal surge starts late, booster vaccination will likely serve to prevent infection best if performed later than the optimal date indicated in this study. Early and late surges are likely associated with factors such as temperature (30, 31), humidity (30), as well as environmentally driven shifts in human behavior (32). Direct research on environmental correlates of surging infection for COVID-19 has been challenging due to pandemic dynamics of SARS-CoV-2, which have been strongly influenced by dramatic spatiotemporal shifts of immune status and of interventions (33, 34). Therefore, accumulation of data regarding environmental correlates of endemic COVID-19 infection would empower the development of predictive seasonal incidence models that could in turn inform dynamic yearly recommendations regarding the timing of booster vaccination.

Our analyses regarding yearly optimal booster date and optimal delay in response to infection are based on a typical healthy adult immune response to vaccination and infection and consequent waning of immunity. However, in modern human populations, immune status is highly heterogeneous (35, 36). Factors such as immune-suppressing therapies (37), HIV (38), nutrition (39, 40), as well as aging (41) can all modulate the immune response. For individuals with compromised immune systems, it may be possible and beneficial to obtain additional booster vaccinations that can stimulate sufficient protection (20). If restricted to a single booster vaccination per year, those with immune-compromising conditions would likely minimize their yearly probability of infection by timing their booster vaccination slightly later than the yearly optimal booster vaccination date determined for a typical healthy adult, ensuring that their lesser, shorter, peak protection spans the time at which peak seasonal infection occurs.

Booster vaccination has tremendous public health potential (20, 42, 43). However, uptake of COVID-19 booster vaccination by low proportions of residents has lowered its public health impact (26). Our results show that appropriate timing of updated booster vaccination can provide a three- to five-fold improvement in the yearly protection provided. Moreover, we address a major gap in knowledge and source of hesitation regarding uptake: we provide previously absent guidance for optimal timing of booster vaccination for individuals who have recently been infected. Perceptions of decreased susceptibility for infection, often stemming from known recent infections, are associated with decreased uptake of boosters (44). For instance, healthcare workers who have been infected recently are unlikely to uptake, and can serve as unintentional drivers of vaccine hesitancy as they are viewed as models for public health responses by much of the general public (45). Explicit guidance on how to adjust booster vaccination timing in response to infection—including in chronically exposed health-care workers—acknowledges individual exposure history for a substantial portion of the population, and provides informed advice regarding their health.

Our analyses are based on Pfizer-BioNTech BNT1262b2 boosters under endemic conditions. However, changes in the efficacy between serially produced booster vaccines or between booster vaccines produced by different manufacturers may alter the relative level of protection provided at peak antibody level and during subsequent waning. Serial or alternate booster vaccines that provoke a lesser antibody response will lead to yearly optimal booster dates and optimal delays following booster infection that are closer to months in which incidence is high. Serial or alternate booster vaccines that provoke a greater antibody response will lead to yearly optimal booster dates and optimal delays following booster infection that precede the months in which incidence is high by a wider margin. Current booster vaccines with high levels of uptake such as those manufactured by Moderna induce antibody responses and protection from infection that is similar to booster vaccines from Pfizer-BioNTech (46). Yearly optimal booster dates and optimal booster vaccination delays following breakthrough infection that are indicated by our analysis should be generally applicable.

In the case of a breakthrough infection, the U.S. Centers for Disease Control currently suggest that individuals may delay their booster uptake by up to three months (9). This three-month delay is appropriate for some mid-year infections. However, this consistency is rather like a stopped clock that is correct at only certain times of the day: if a breakthrough infection occurred shortly after booster vaccination on the yearly optimal booster vaccination date, it is not optimal to delay the next booster at all beyond the next yearly optimal booster vaccination date. If a breakthrough infection occurred much later in the year following booster vaccination on the yearly optimal booster vaccination date—a more likely scenario—then it is optimal to delay the booster from the yearly optimal booster vaccination date to a much later date. Additionally, just as the yearly optimal booster vaccination date is moderately affected by the seasonal incidence patterns inherent to the geographic location, the optimal delay is moderately affected by geographic variation in incidence as well.

These results for COVID-19 provide the first continuous assessment of risk of infection with respect to annual booster vaccination for both people whose most recent exposure was a scheduled booster, and for people whose most recent exposure was a breakthrough infection. For respiratory viruses with available vaccines, immunity wanes following vaccination increasing the risk of breakthrough infection (47–49). Robust analyses of optimal vaccine timing incorporating immuno-epidemiological inference (50) regarding numerous diseases including influenza and RSV are warranted, empowered by requisite collection of long-term longitudinal immunological cohort data from relevant individuals and long-term seasonal incidence data from diverse geographic locales. Together, immuno-epidemiological inference based on long-term infection data can play a substantial role in increasing the efficacy of booster vaccination, curtailing morbidity and mortality due to respiratory infectious disease.

## Data Availability

All data, Mathematica notebooks, imputed monthly proportions, and code underlying this study are publicly available on Zenodo: DOI:10.5281/zenodo.11209811.

https://zenodo.org/records/11209811

## Authors contributions

JPT, HBH, and AD conceived the project and designed the study; HBH performed formal analyses with guidance from JPT and AD; HBH, JPT, and AD designed data visualizations; HBH implemented data visualizations; AD and JPT wrote the manuscript; and all authors reviewed the manuscript before submission. JPT and AD were responsible for the decision to submit the manuscript. All authors had full access to all the data in the study and had final responsibility for the decision to submit for publication.

## Declarations of interest

None.

## Funding

National Science Foundation of the United States of America RAPID 2031204 (JPT and AD), NSF Expeditions CCF 1918784 (JPT and APG), and support from the University of North Carolina, Charlotte to AD.

## Supplementary Information

Supplementary Information is available for this paper.

Correspondence and requests for materials should be addressed to Jeffrey P. Townsend.

## Notes

### Competing Interest Statement

The authors have declared no competing interest.

