## Supplemental Materials for "Optimal annual COVID-19 vaccine boosting dates following previous booster vaccination or breakthrough infection"

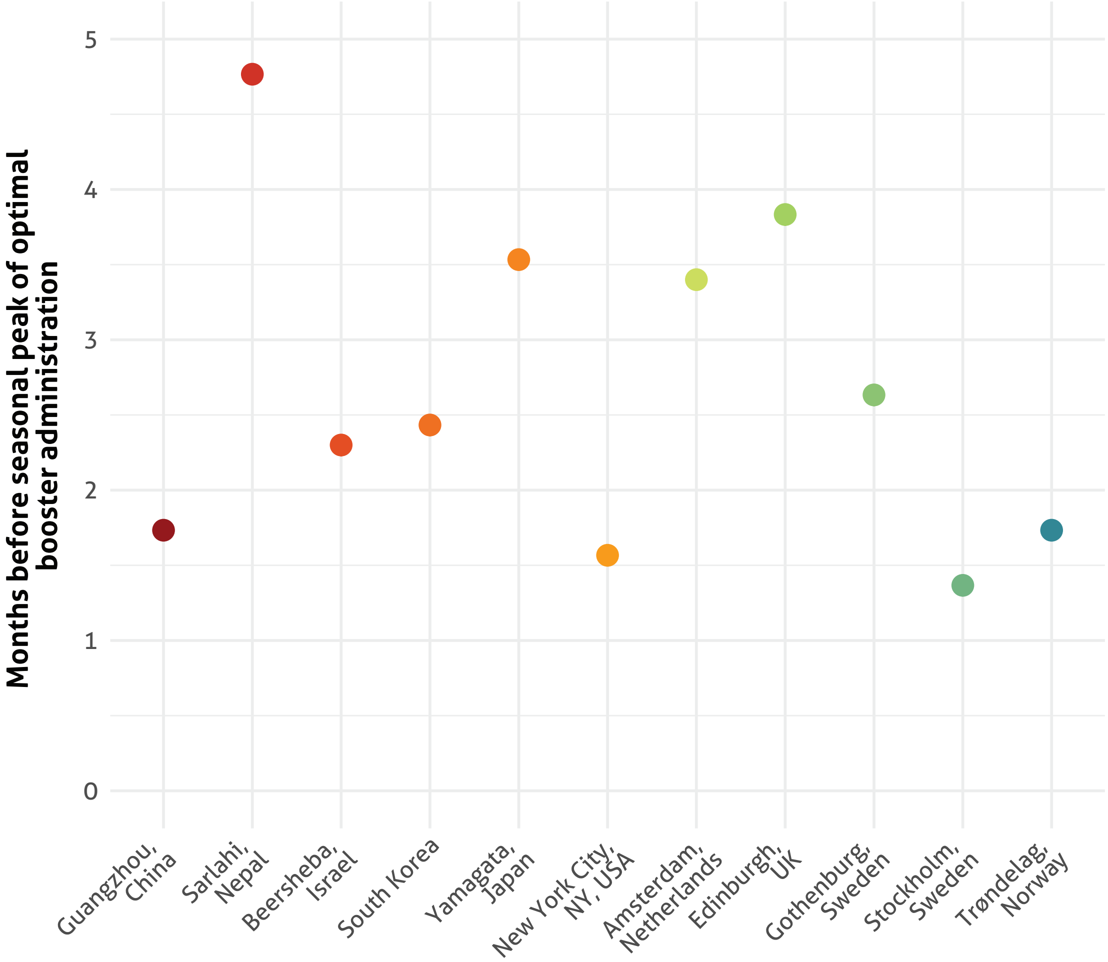


**Supplementary Figure 1.** Month of optimal booster date versus the month exhibiting the highest yearly seasonal incidence in 11 locations across the Northern Hemisphere

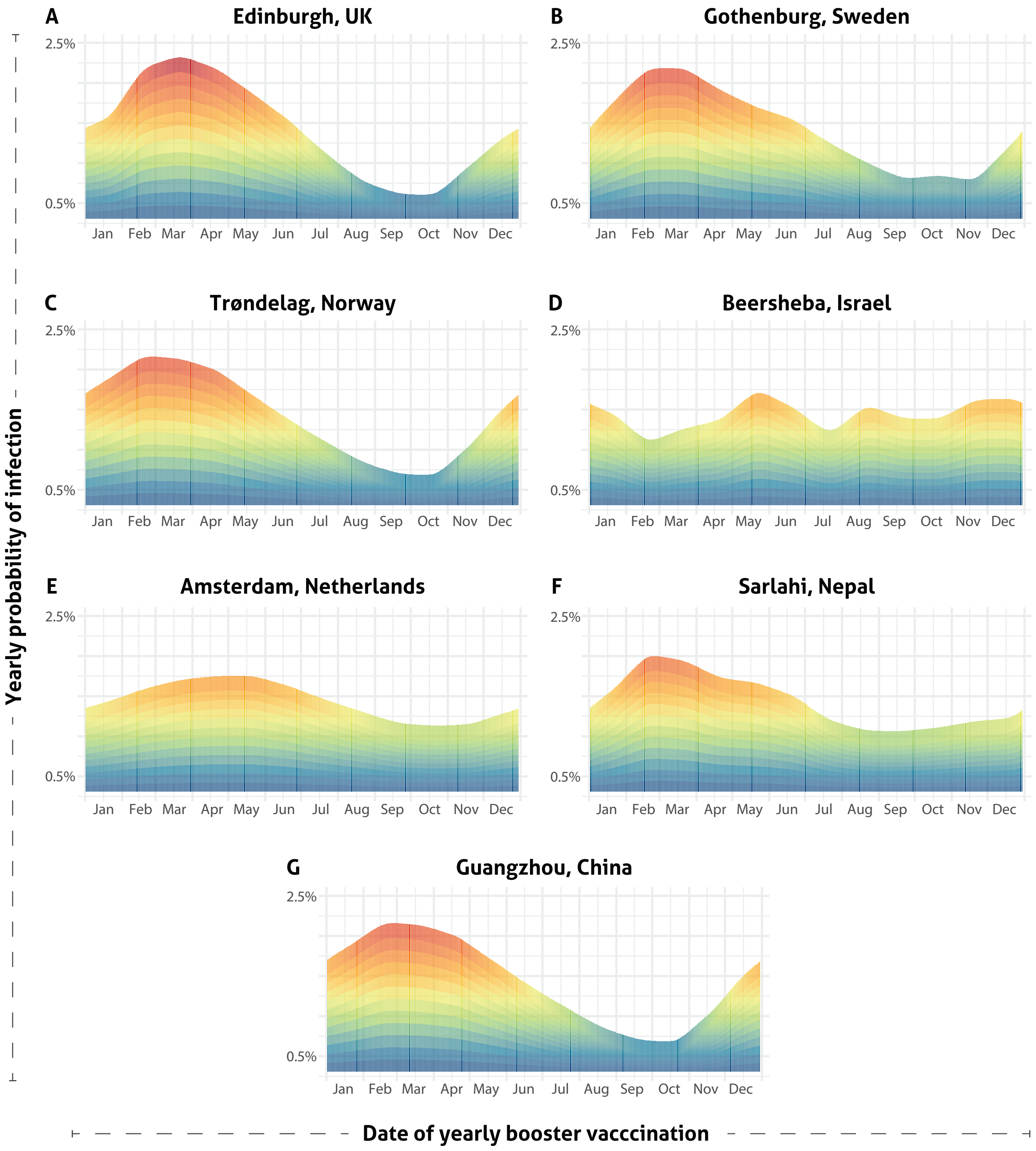


**Supplementary Figure 2.** Yearly probability of infection (red: high to blue: low) based on yearly date of booster vaccination in 7 additional locations across the Northern Hemisphere.

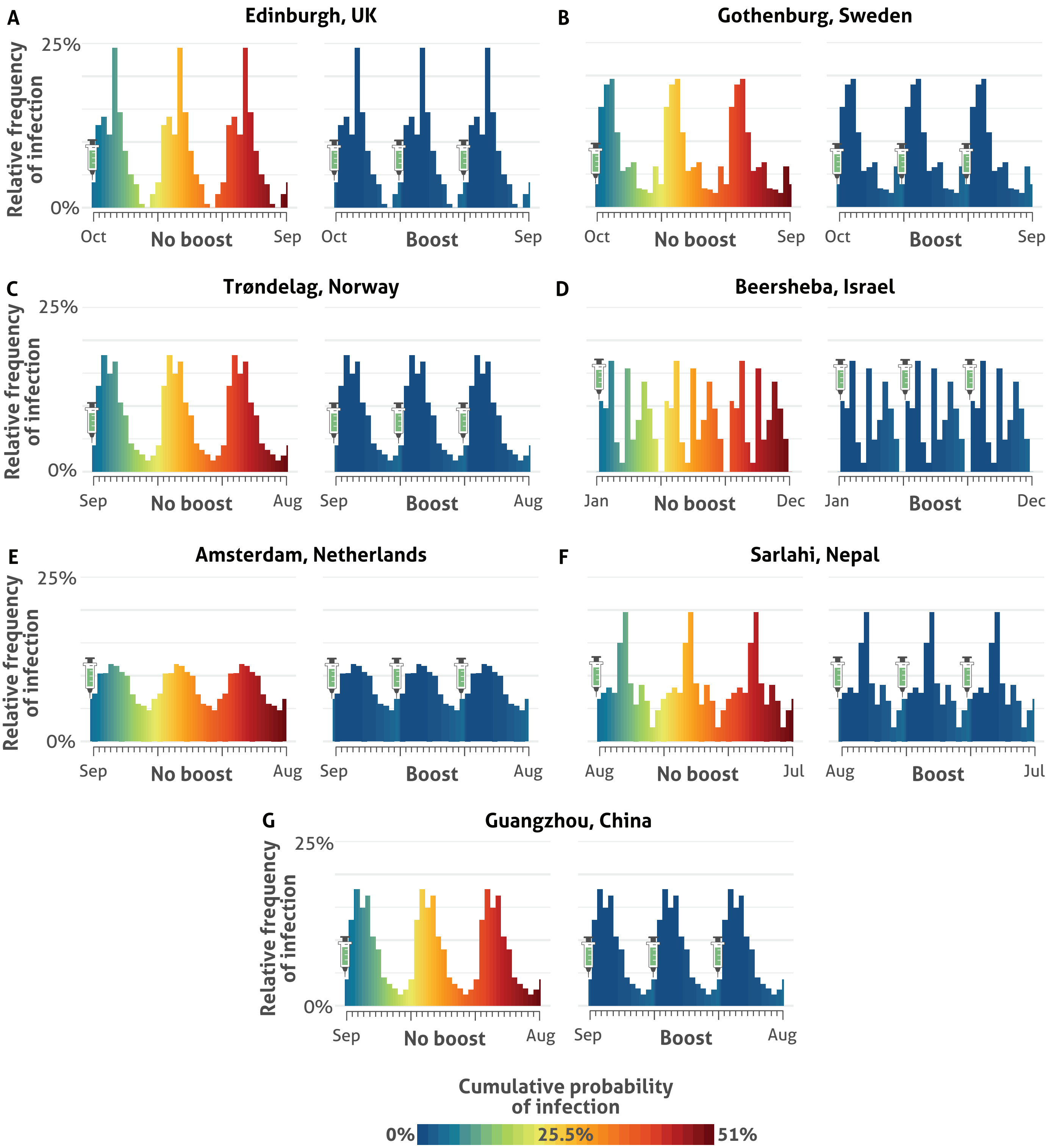


**Supplementary Figure 3.** Cumulative probabilities of infection—indicated by the blue / green / yellow / orange / red color gradient—in the year after booster vaccination on the optimal date and in the two following years without the yearly booster vaccination, in 7 additional locations across the Northern Hemisphere. The heights of the bars reflect projections of seasonal population-level incidence patterns under endemic conditions without incorporating a population-wide effect of booster vaccination, i.e. assuming that low yearly global uptake (Jacobs et al. 2023) remains unchanged.

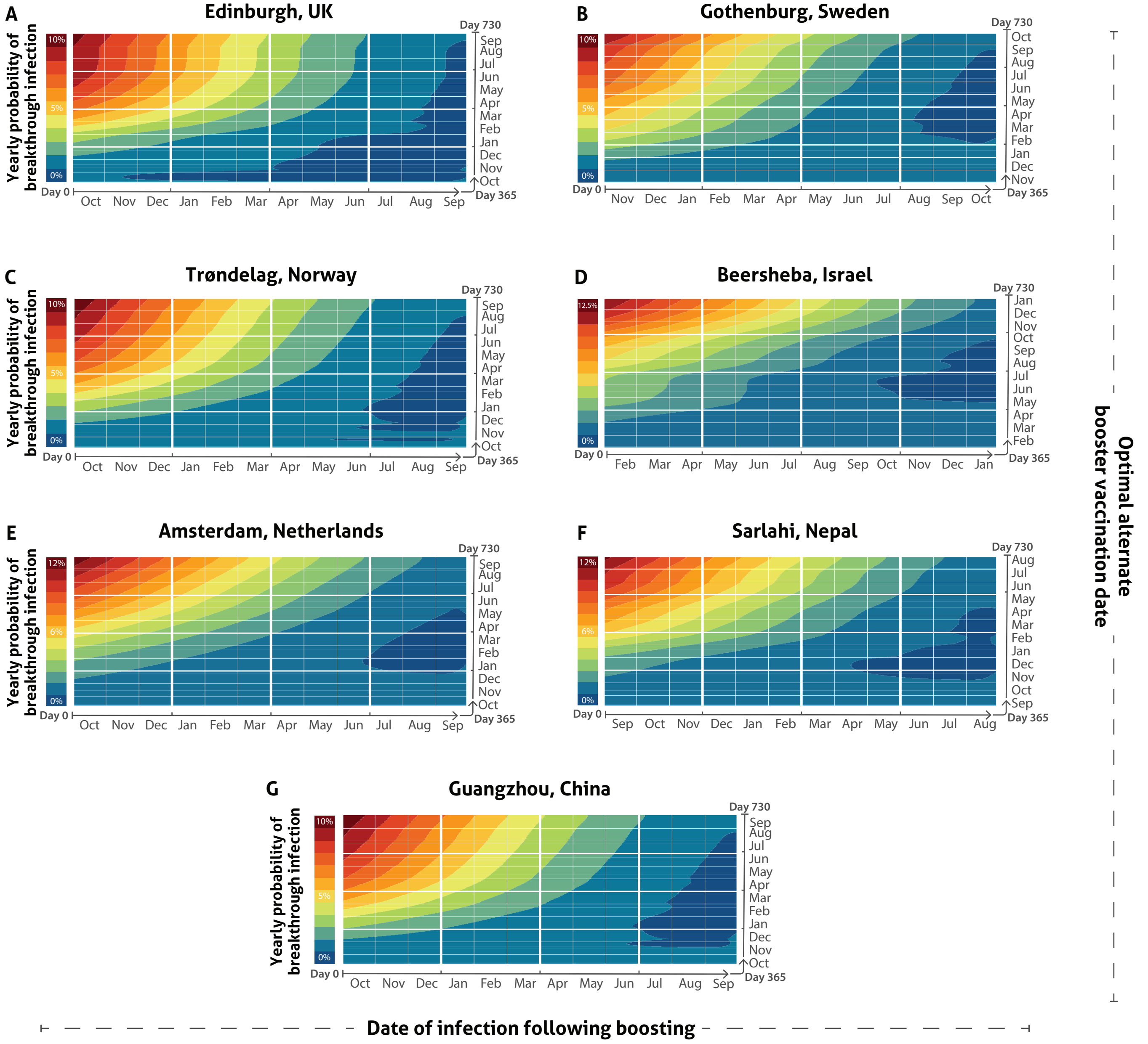


**Supplementary Figure 4.** Probability of infection upon delaying booster vaccination to each date of the year, in response to a breakthrough infection occurring in the interval between optimal booster vaccination dates in 7 additional locations across the Northern Hemisphere. Day 365 is the approximate optimal yearly booster vaccination date. Infection will delay the optimal booster vaccination date beyond day 365 to a greater and greater degree as the interval between the yearly optimal booster date and the date of infection increases.
